# Myocardial Infarction across COVID-19 Pandemic Phases: Insights from the Veterans Health Affairs System

**DOI:** 10.1101/2023.03.07.23286963

**Authors:** Celina Yong, Laura Graham, Tariku J. Beyene, Shirin Sadri, Juliette Hong, Tom Burdon, William F. Fearon, Steven M. Asch, Mintu Turakhia, Paul Heidenreich

## Abstract

**Background:** Cardiovascular procedural treatments were deferred at scale during the COVID-19 pandemic, with unclear impact on patients presenting with Non-ST Elevation Myocardial Infarction (NSTEMI).

**Methods:** In a retrospective cohort study of all patients diagnosed with NSTEMI in the U.S. Veterans Affairs Healthcare System from 1/1/19 to 10/30/22 (n=67,125), procedural treatments and outcomes were compared between the pre-pandemic period and six unique pandemic phases (1: Acute phase, 2: Community spread, 3: First Peak, 4: Post-Vaccine, 5. Second Peak, 6. Recovery). Multivariable regression analysis was performed to assess association between pandemic phases and 30-day mortality.

**Results:** NSTEMI volumes dropped significantly with the pandemic onset (62.7% of pre-pandemic peak) and did not revert to pre-pandemic levels in subsequent phases, even after vaccine availability. Percutaneous coronary intervention (PCI) and/or coronary artery bypass grafting (CABG) volumes declined proportionally. Compared to the pre-pandemic period, NSTEMI patients experienced higher 30-day mortality during Phase 2 and 3, even after adjustment for COVID-19 positive status, demographics, baseline comorbidities, and receipt of procedural treatment (adjusted OR for Phase 2-3 combined: 1.26 [95% CI 1.13-1.43], p<0.01). Patients receiving VA-paid community care had a higher adjusted risk of 30-day mortality compared to those at VA hospitals across all six pandemic phases.

**Conclusions:** Higher mortality after NSTEMI occurred during the initial spread and first peak of the pandemic, but resolved before the second, higher peak – suggesting effective adaptation of care delivery but a costly delay to implementation. Investigation into the vulnerabilities of the early pandemic spread are vital to informing future resource-constrained practices.

**Clinical Perspective:** *What is New?:*

- After the initial significant decline in NSTEMI presentations during the acute phase of the pandemic, volumes of NSTEMI presentations and procedural treatment have not reverted to pre-pandemic levels despite widespread availability of vaccines in the Veterans Health Administration.
- Compared to the pre-pandemic period, 30-day mortality after NSTEMI increased during the initial spread and first pandemic peak (Phases 2 and 3) -- but resolved before the subsequent highest pandemic peak of Phase 5 -- suggesting a delay to implementation of adapted systems of cardiovascular care.
- The increased mortality was not significantly mediated by the decline in procedural volumes, suggesting appropriate triage of procedural care during the pandemic.

*What are the Clinical Implications?:*

- The COVID-19 pandemic appears to have had a lasting impact on health-seeking behaviors among NSTEMI patients, with unclear long-term effects of this increased threshold to obtain cardiovascular care.
- Investigation into the vulnerabilities that occurred during initial phases of the pandemic are urgently needed to inform ongoing and future resource-constrained practices.

## Introduction

In efforts to curb the spread of the novel coronavirus (SARS-CoV-2) and direct resources to address COVID-19 pandemic needs, Veterans Affairs (VA) hospitals nationwide received unprecedented mandates starting in March 2020 to postpone all elective cardiovascular procedures, while permitting only urgent, life-threatening ones.^1,2^ Given that PCI for myocardial infarction (MI) has been proven to reduce major adverse cardiac events,^3,4^ impacts of this procedural triage have been unclear.^5,6,7,8^ The evolving pandemic has unintentionally offered a natural experiment to demonstrate the transition from large scale de-implementation of elective cardiovascular procedural care through risk stratification to recovery. As the largest integrated healthcare system in the United States, the Veterans Affairs Healthcare System offers a unique opportunity to understand the impact of these adaptations. In this study, we examine how the evolving treatment paradigm during the prolonged pandemic has impacted patients with Non-ST-Elevation Myocardial Infarction (NSTEMI).

## Methods

We performed a retrospective cohort study of all patients diagnosed with NSTEMI between 1/1/2019 to 10/30/22 who received inpatient care at a VA hospital or paid for by the Veterans Health Administration (VHA). Patient cohort selection is shown in **Supplemental Figure 1**. The VHA provides care to over 9 million Veterans across the United States at more than 1200 healthcare facilities, including 171 VA Medical Centers.^9^

**Figure 1.**
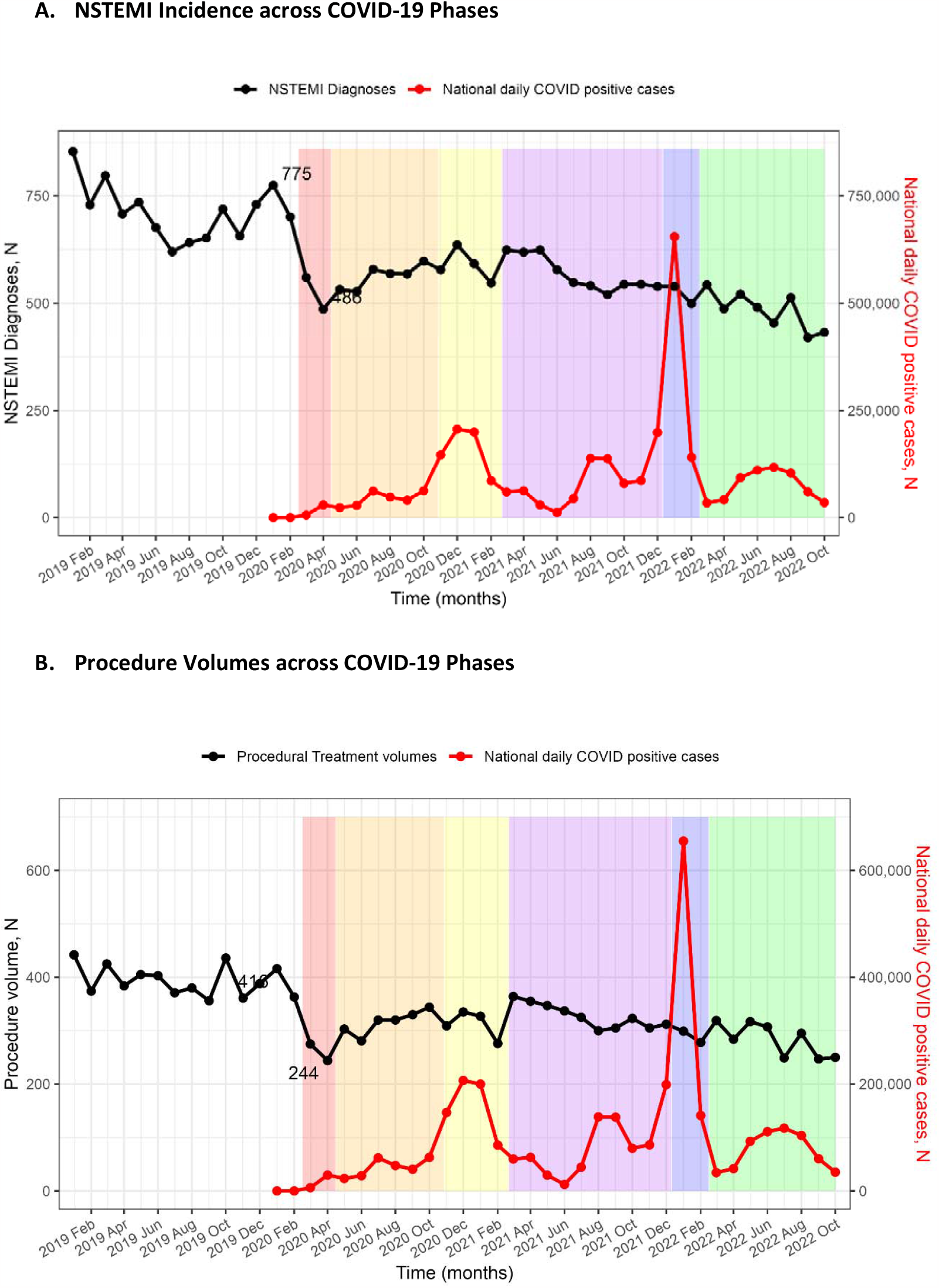
NSTEMI Diagnoses and Procedural Treatment at VA Facilities by COVID-19 Phase. A. **NSTEMI Incidence across COVID-19 Phases**. Volumes of NSTEMI presentations declined in Phase 1, which did not recover to pre-COVID levels in subsequent phases. B. **Procedure Volumes across COVID-19 Phases**. Procedural treatments (angiogram/PCI/CABG) for NSTEMI underwent a decline in Phase 1, followed by steady volumes over subsequent phases.

Patients requiring inpatient care for NSTEMI were identified by International Classification of Diseases, Tenth Revision, Clinical Modification (ICD-10-CM) diagnosis codes (NSTEMI: I21.4) from the VA Corporate Data Warehouse (CDW). ^10,11^ Additional covariates included patient demographics, socioeconomic characteristics, and comorbidity burden, including Charlson Comorbidity Index. Patient geographic information (ZIP code and distance to medical centers) was obtained from the VA Planning Systems Support Group (PSSG) data. U.S. Census data based on patient zip code was used to include neighborhood education level, and income status. Procedures (angiograms, percutaneous coronary intervention, and coronary artery bypass grafting) performed within 30 days of incident diagnosis were identified from the VA CDW by ICD-10 and CPT procedure codes, as well as from VA CART-PCI data (**Supplemental Table 1**). Mortality was obtained from the VA CDW Vital Status domain.

**Table 1.** Baseline Patient Characteristics across COVID-19 Phases.

|  | COVID Phase |  |  |  |  |  |  | P-value |
| --- | --- | --- | --- | --- | --- | --- | --- | --- |
|  | Pre-COVID | Phase 1 | Phase 2 | Phase 3 | Phase 4 | Phase 5 | Phase 6 |  |
|  | 1/1/2019 –<br>15/2/2020 | 16/2/2020 –<br>15/4/2020 | 16/4/2020 –<br>27/11/2020 | 28/11/2020 –<br>202/2021 | 20/2/2021 –<br>10/11/2021 | 11/11/2021 –<br>15/2/2022 | 16/2/2022 –<br>30/10/2022 |  |
| Characteristics | (N=9682) | (N=1097) | (N=4155) | (N=1682) | (N=5470) | (N=1183) | (N=4077) |  |
| **Age, yrs, mean(SD)** | 71.4 (10.69) | 71.4 (11.11) | 71.1 (10.75) | 72.0 (11.17) | 71.8 (10.96) | 72.4 (11.17) | 72.3 (10.74) | <0.01<sup>2</sup> |
| **Gender, n (%)** |  |  |  |  |  |  |  | <0.01<sup>1</sup> |
| Female | 265 (2.7%) | 48 (4.4%) | 124 (3.0%) | 46 (2.7%) | 194 (3.5%) | 42 (3.6%) | 141 (3.5%) |  |
| Male | 9417 (97.3%) | 1049 (95.6%) | 4031 (97.0%) | 1636 (97.3%) | 5276 (96.5%) | 1141 (96.4%) | 3936 (96.5%) |  |
| **Race/Ethnicity, n (%)** |  |  |  |  |  |  |  | 0.013<sup>1</sup> |
| White | 6247 (68.5%) | 675 (66.0%) | 2636 (67.3%) | 1061 (66.7%) | 3447 (67.1%) | 743 (67.2%) | 2490 (65.2%) |  |
| Black | 1994 (21.9%) | 233 (22.8%) | 896 (22.9%) | 368 (23.1%) | 1158 (22.5%) | 253 (22.9%) | 914 (23.9%) |  |
| Hispanic | 697 (7.6%) | 83 (8.1%) | 303 (7.7%) | 120 (7.5%) | 383 (7.5%) | 75 (6.8%) | 298 (7.8%) |  |
| Other | 187 (2.0%) | 32 (3.1%) | 84 (2.1%) | 41 (2.6%) | 150 (2.9%) | 35 (3.2%) | 118 (3.1%) |  |
| **Marital Status, n (%)** |  |  |  |  |  |  |  | 0.08<sup>1</sup> |
| Married | 4435 (45.9%) | 504 (46.4%) | 1905 (46.0%) | 771 (46.0%) | 2572 (47.3%) | 545 (46.5%) | 1987 (48.9%) |  |
| Separated | 4211 (43.6%) | 461 (42.4%) | 1786 (43.2%) | 731 (43.6%) | 2268 (41.7%) | 491 (41.9%) | 1623 (40.0%) |  |
| Single | 1016 (10.5%) | 121 (11.1%) | 448 (10.8%) | 173 (10.3%) | 599 (11.0%) | 135 (11.5%) | 450 (11.1%) |  |
| **Rurality by, n (%)** |  |  |  |  |  |  |  | 0.04<sup>1</sup> |
| Highly rural | 279 (2.9%) | 30 (2.8%) | 148 (3.6%) | 43 (2.6%) | 161 (3.0%) | 29 (2.5%) | 127 (3.2%) |  |
| Rural | 2591 (26.9%) | 257 (23.9%) | 1056 (25.8%) | 412 (24.9%) | 1395 (26.1%) | 312 (26.9%) | 965 (24.4%) |  |
| Urban | 6770 (70.2%) | 788 (73.3%) | 2891 (70.6%) | 1201 (72.5%) | 3797 (70.9%) | 819 (70.6%) | 2865 (72.4%) |  |
| **Education Level, n %** |  |  |  |  |  |  |  | <0.01<sup>1</sup> |
| <25% of HS or less | 8022 (82.9%) | 890 (81.1%) | 3495 (84.1%) | 1397 (83.1%) | 4646 (84.9%) | 983 (83.1%) | 3484 (85.5%) |  |
| >=25% of HS or less | 1660 (17.1%) | 207 (18.9%) | 660 (15.9%) | 285 (16.9%) | 824 (15.1%) | 200 (16.9%) | 593 (14.5%) |  |
| **Income, % (n)** |  |  |  |  |  |  |  | 0.97<sup>1</sup> |
| <=25% 75K or less | 7935 (82.0%) | 904 (82.4%) | 3407 (82.0%) | 1384 (82.3%) | 4501 (82.3%) | 977 (82.6%) | 3322 (81.5%) |  |
| >25% 75K or less | 1747 (18.0%) | 193 (17.6%) | 748 (18.0%) | 298 (17.7%) | 969 (17.7%) | 206 (17.4%) | 755 (18.5%) |  |

| **Comorbidities** |  |  |  |  |  |  |  |  |
| --- | --- | --- | --- | --- | --- | --- | --- | --- |
| CCI, mean (SD) | 5.7 (3.22) | 5.8 (3.28) | 5.6 (3.23) | 5.5 (3.14) | 5.6 (3.24) | 5.5 (3.16) | 5.6 (3.28) | 0.01<sup>2</sup> |
| COVID pos, n (%) | 0 (0.0%) | 26 (2.4%) | 147 (3.5%) | 139 (8.3%) | 165 (3.0%) | 134 (11.3%) | 174 (4.3%) | <.01<sup>1</sup> |
| CHF, n (%) | 5312 (54.9%) | 588 (53.6%) | 2252 (54.2%) | 890 (52.9%) | 2938 (53.7%) | 626 (52.9%) | 2122 (52.1%) | 0.17<sup>1</sup> |
| PVD, n (%) | 3241 (33.5%) | 361 (32.9%) | 1287 (31.0%) | 521 (31.0%) | 1690 (30.9%) | 355 (30.0%) | 1265 (31.1%) | <0.01<sup>1</sup> |
| CVD, n (%) | 2109 (21.8%) | 250 (22.8%) | 850 (20.5%) | 359 (21.3%) | 1201 (22.0%) | 263 (22.2%) | 879 (21.6%) | 0.54<sup>1</sup> |
| COPD, n (%) | 3726 (38.5%) | 433 (39.5%) | 1482 (35.7%) | 589 (35.0%) | 1940 (35.5%) | 401 (33.9%) | 1433 (35.2%) | <.01<sup>1</sup> |
| Diabetes WOC, n (%) | 5212 (53.8%) | 570 (52.0%) | 2173 (52.3%) | 873 (51.9%) | 2923 (53.4%) | 613 (51.8%) | 2117 (52.0%) | 0.26<sup>1</sup> |
| Diabetes WC, n (%) | 3955 (40.9%) | 445 (40.6%) | 1658 (39.9%) | 629 (37.4%) | 2228 (40.7%) | 459 (38.8%) | 1638 (40.2%) | 0.17<sup>1</sup> |
| Renal Disease, n (%) | 4013 (41.5%) | 480 (43.8%) | 1713 (41.2%) | 673 (40.0%) | 2224 (40.7%) | 506 (42.8%) | 1664 (40.8%) | 0.39<sup>1</sup> |
| Cancer, n (%) | 1740 (18.0%) | 208 (19.0%) | 781 (18.8%) | 307 (18.3%) | 1016 (18.6%) | 227 (19.2%) | 819 (20.1%) | 0.16<sup>1</sup> |
| Metastatic Carcinoma, n (%) | 391 (4.0%) | 51 (4.6%) | 172 (4.1%) | 61 (3.6%) | 205 (3.7%) | 38 (3.2%) | 189 (4.6%) | 0.17<sup>1</sup> |

<sup>1</sup>Chi-Square p-value; <sup>2</sup>Kruskal-Wallis p-value; CCI= Charlson Comorbidity Index, CHF= Congestive Heart Failure, PVD=Peripheral Vascular Disease, CVD= Cerebrovascular Disease, COPD= Chronic Obstructive Pulmonary Disease, Diabetes WC= Diabetes with complications Diabetes WOC= Diabetes without complications.

Patients were categorized into one of six COVID phases according to the date of their first NSTEMI diagnosis, with phase dates defined by epidemiological trends in the United States:^12,13^

**Pre-COVID Phase** was defined as the year prior to the COVID-19 pandemic (01/01/2019-02/15/2020).

**Phase 1** (“Acute Phase”, 02/16/20-04/15/20) was defined as the two-month period surrounding the initial nadir of patient NSTEMI volumes due to the pandemic, which included the initial period of national VA directives to defer non-essential procedures. **Phase 2** (“Community Spread”, 04/16/2020-10/27/2020) was the initial period of rapidly rising COVID infections in the United States.

**Phase 3** (“First Peak”, 10/28/2020-02/20/2021) was the first epidemiologic peak of COVID infections in the United States.

**Phase 4** (“ Post-Vaccine”, 02/21/2021-12/10/2021) represented the period after which all Veterans who desired vaccination should have received it. By this time, most catheterization labs had resumed routine practices with restrictions limited to pre-procedure COVID-19 testing requirements.

**Phase 5** (“Second Peak”, 12/11/2021-02/15/2022) represented the highest epidemiologic peak of COVID infections in the United States, largely due to new variants.

**Phase 6** (“Recovery Phase”, 02/16/2022-10/30/2022) represented the endemic period after removal of indoor COVID-19 restrictions in most states.

Among all NSTEMI patients presenting to VA PCI-capable hospitals, patient demographics and comorbidities (within one year prior to first NSTEMI diagnosis) were compared across COVID phases using Chi-square for categorical variables and ANOVA for continuous variables. Trend line visualization was used to examine average changes in diagnoses and procedural volumes across COVID phases in comparison with national daily average COVID-19 positive cases. Multivariable logistic regression was performed to evaluate 30-day mortality by COVID phase (pre-COVID as reference). Model covariate selection was performed with LASSO (SAS procedure hpgenselect using the Schwarz Bayes Criterion (SBC) with lambda = 0.8 and 25 steps), which yielded inclusion of age, Charlson Comorbidity Index (CCI), and COVID-19 positive test status at the time of admission in the final adjusted models. We also performed additional adjustment for receipt of procedural treatment to assess its potential contribution to the outcome. Kaplan Meier curves were used to compare mortality over time across COVID-19 phases. Unlike private health insurance, VA enrollment is typically stable over the life of the patient without disenrollment. To evaluate stability of patient enrollment over time in our cohort, a separate analysis of monthly outpatient prescription volumes was performed, showing no significant changes over time (with the exception of a brief decline in Phase 1 that returned to baseline by Phase 2). Multivariate logistic regression was also used to compare 30-day mortality among patients presenting with NSTEMI to VA vs. Non-VA facilities across COVID phases, with adjustment for demographics and comorbidities. Wald Chi-Squared test and odds ratios were used to compare likelihood of 30-day mortality between VA and Non-VA (Fee-Basis) care patients in pre- vs. post-COVID periods. This was a complete case analysis, with no missing values for the adjusted covariates (age, CCI, COVID positive test status, or receipt of procedure). Two sided tests were used in all scenarios. This study was approved by the Stanford Institutional Review Board. Analyses were conducted in SAS v9.4.

## Results

A total of 67,125 Veterans were coded with a new NSTEMI from 1/1/19 to 10/30/2022, of whom 27,346 presented to a PCI-capable VA hospital.

Over progressive pandemic phases, the mean age of patients presenting with NSTEMI increased (**Table 1**). Patient race/ethnicity also varied by phase (p=0.01), with the highest proportion of Black patients presenting with NSTEMI during Phase 3 (23.2%) and the highest proportion of Hispanic patients presenting with NSTEMI in Phase 1 (8.1%). Rural patients and those with lower education levels had lower NSTEMI presentations during the acute phase (Phase 1) (p=0.04 and p<0.01 respectively, across all phases). The highest proportion of NSTEMI patients with COVID-19 infection presented during the second peak (11.3% in Phase 5, p<0.01 across all phases).

NSTEMI presentations during the acute phase (Phase 1) of the pandemic reached a nadir during the month of April 2020, with 486 NSTEMIs nationally that month, representing 62.7%, of volume at the pre-pandemic peak (week of January 1, 2020, **Figure 1A**). After the initial nadir, there was a slight uptrend in NSTEMI presentations in early Phase 2, which was followed by an overall downwards trend over time that did not revert to pre-pandemic levels even after vaccine roll-out that began in December 2020 (during Phase 3). **Figure 1A** demonstrates that the seasonal variation in NSTEMI volumes during the pre-pandemic year was no longer observed during the subsequent pandemic years. Evaluation of monthly national outpatient prescription refills showed stability over time (aside from a brief decline in Phase 1), suggesting negligible contribution to the decline in NSTEMI volumes.

Procedures performed to diagnose and treat NSTEMI (angiogram, PCI, CABG) also declined during Phase 1. The nadir of 244 procedures among NSTEMI patients during the month of April 2020 represents 58.7% of the pre-pandemic peak (week of January 1, 2020, **Figure 1B**). While the absolute numbers declined, the proportion of NSTEMI patients receiving PCI and/or CABG did not significantly change (33.6% pre-COVID vs. 34.0% post-COVID, p=0.51, volumes by phase in **Supplemental Table 2**). When angiogram was added to the list of procedures, there was still no significant difference in probability of receiving a procedure (55.6% pre-COVID vs. 56.8% post-COVID, p=0.09). Logistic regression also showed no significant relationship between procedural volumes and all COVID phases combined (likelihood of procedural treatment compared to pre-COVID: adjusted OR 1.05 [95% CI 0.99-1.10], p=0.09). However, analysis of each individual phase showed a significantly lower likelihood of receiving a procedure during Phase 1 (adjusted OR 0.77 [95% CI 0.77-0.64], p=0.002), and a significantly higher likelihood of receiving a procedure during Phase 6 (adjusted OR 1.18 [95% CI 1.05-1.33], p=0.0005, **Supplemental Table 3** for other non-significant phase results).

Analysis of survival plots revealed that NSTEMI mortality was higher in phases 2, 3, and 5 compared to the pre-pandemic period (**Figure 2**, Phase 2: OR 1.13([95% CI 1.04, 1.23], p<0.01); Phase 3 OR 1.17([95% CI 1.04, 1.31], p<0.01); Phase 5 OR 1.15([95% CI 1.01, 1.31], p=0.04). There were no significant differences in the other phases.

**Figure 2.**
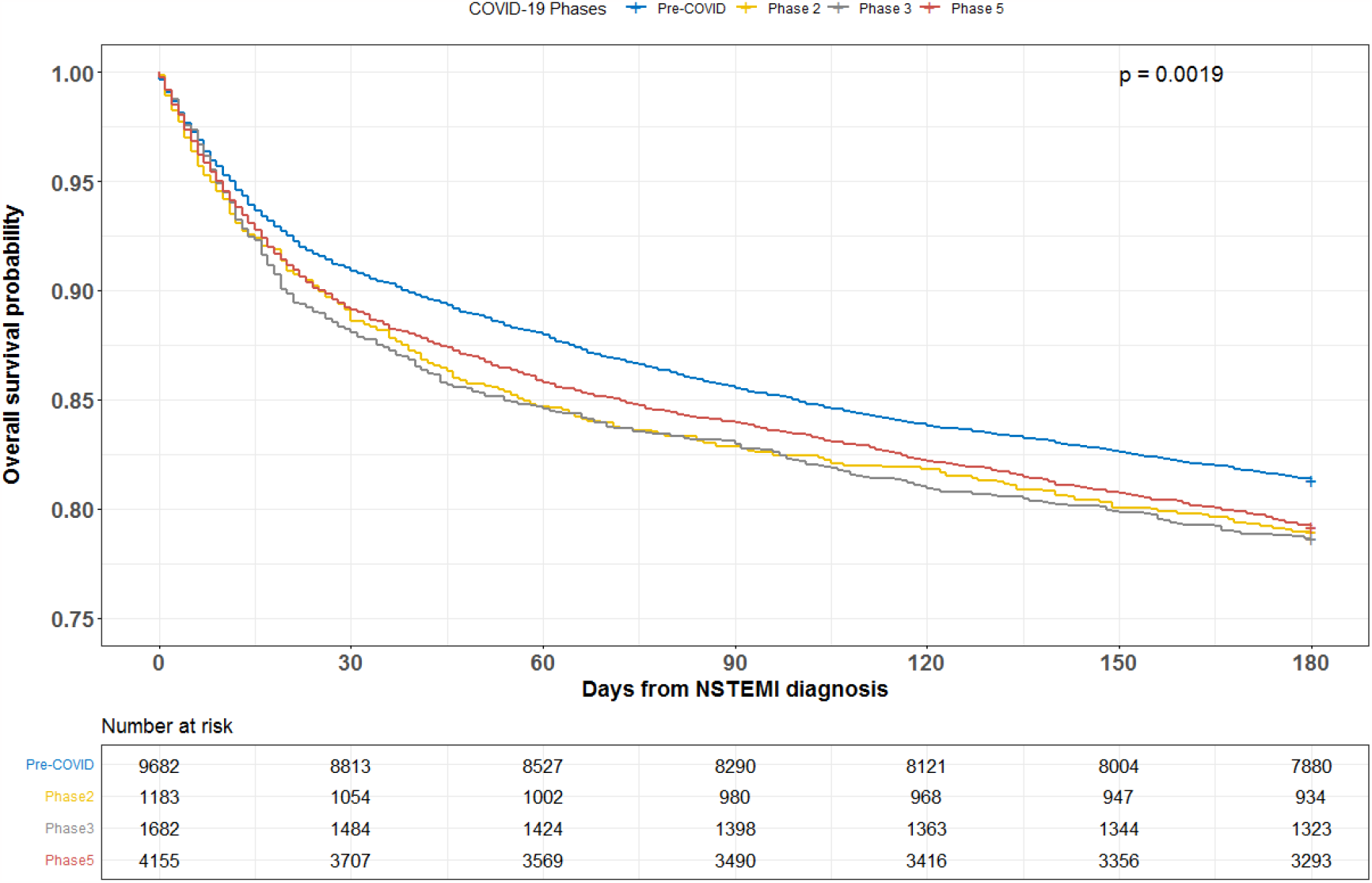
Kaplan Meier Survival Plot by COVID-19 Phases. Phases 2, 3, and 5 showed significantly higher mortality compared to pre-COVID (non-significant phases not shown).

Unadjusted logistic regression analysis revealed that from Phase 2 (community spread) up to Phase 5 (second peak), NSTEMI patients had an average higher 30-day mortality compared to pre-pandemic. After adjustment for demographics, baseline comorbidities, COVID-19 positive infection status, and receipt of procedural treatment, these higher mortality findings remained significant for Phases 2 and 3, but not for the other phases (**Figure 3**, unadjusted results in **Supplemental Figure 2**). Note that by Phase 6 (recovery), the risk of mortality reversed to be lower than the pre-pandemic period. Among the 76 catheterization sites represented, procedural volume was not a significant predictor in the fully adjusted NSTEMI model (p=0.16).

**Figure 3.**
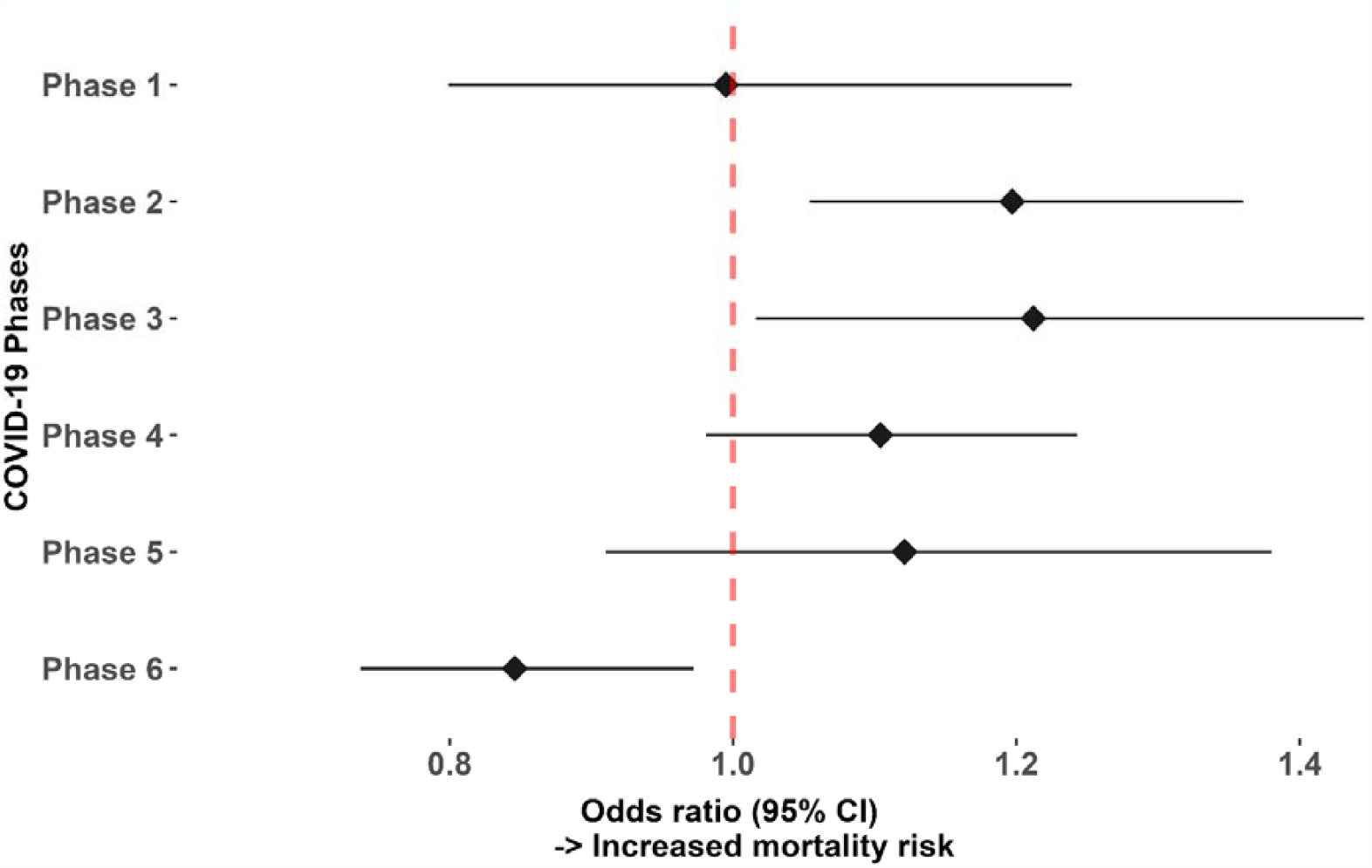
Adjusted 30-day Mortality at VA Facilities by COVID-19 Phase. (reference: pre-COVID phase). Compared to the pre-COVID phase, higher mortality among NSTEMI patients was found in Phase 2 and 3 (adjusted for baseline demographics, comorbidities, COVID status on admission, and receipt of PCI).

There were 20,786 Veteran patients who presented to community care facilities with NSTEMI over the study period (“Fee-Basis” care paid for by VA). The proportion of NSTEMI patients with 30-day mortality among Fee-Basis patients increased over the course of the pandemic, reaching a peak during Phase 3 (**Figure 4A**). Compared to those treated at VA facilities, the risk of 30-day mortality among those treated at non-VA facilities was higher across all pandemic phases. However, after adjustment for demographics and comorbidities, it remained higher only during Phase 3, 4, and 6 (**Figure 4B**, unadjusted results in **Supplemental Figure 3**).

**Figure 4.**
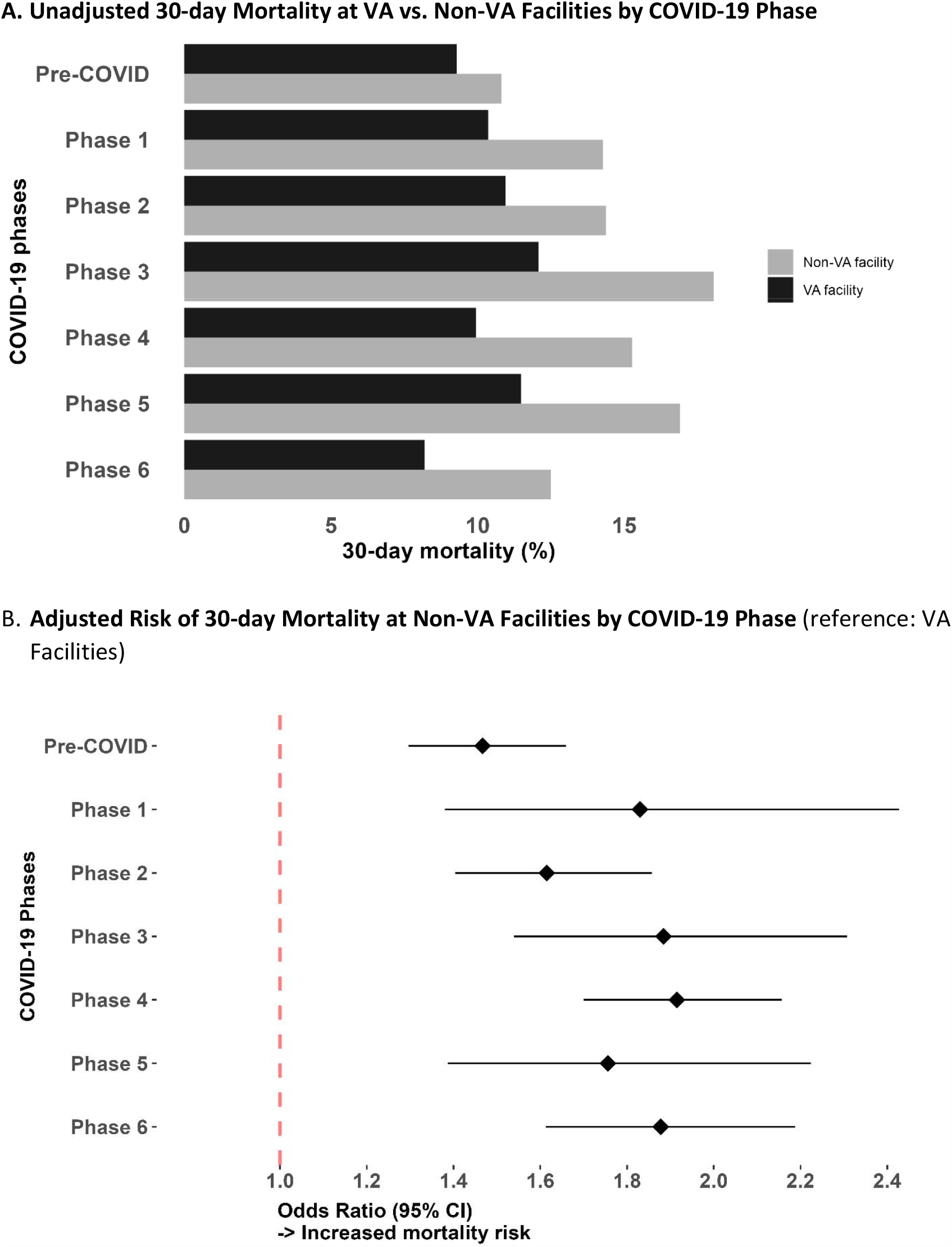
Comparison of 30-day NSTEMI Mortality at VA vs. Non-VA Facilities across COVID-19 Pandemic Phases. **A. Unadjusted 30-day NSTEMI Mortality at VA vs. Non-VA Facilities by COVID-19 Pandemic Phase**. Non-VA facility use peaked during Phases 3 and 5, corresponding to the first and second pandemic peaks. **B. Adjusted Risk of 30-day NSTEMI Mortality at Non-VA Facilities by COVID-19 Phase** (reference: VA facilities). Fee-Basis care outside the VA was associated with higher mortality during multiple pandemic phases compared to treatment within the VA (adjustment for demographics and comorbidities).

## Discussion

In this study, we report longitudinal changes in presentation, treatments, and outcomes among patients with NSTEMI over multiple phases of the COVID-19 pandemic in the Veterans Affairs Healthcare System. We present three key novel findings: 1. The availability of vaccines to Veterans did not significantly change the downtrending trajectory in volumes of patients presenting with NSTEMI, which have not reverted to pre-pandemic levels despite resolving COVID-19 infection burden; 2. Patients presenting with NSTEMI in Phase 2 and 3 had higher 30-day mortality, suggesting vulnerabilities during the initial spread and peak of COVID-19 infections that did not persist despite later, higher COVID infection rates; 3. The increased mortality was not mediated by a decline in procedural volumes, suggesting appropriate triage of procedural care during the pandemic. Notably, over the course of the pandemic, a higher proportion of patients sought non-VA care, which was associated with higher mortality during certain phases of the pandemic even after risk adjustment.

In the VHA, the volume of NSTEMI presentations during Phase 1 of the pandemic dropped dramatically – as has been reported in other non-COVID related disease states.^14^ Given the variability of severity and symptoms within the NSTEMI diagnosis, patients who could tolerate milder symptoms may have stayed home out of fear of contagion. Steady outpatient prescription refills over time in the national VA system argue against competing risk of death due to COVID-19 infection as a major contribution to the decline. The pandemic induced exacerbation of pre-existing access problems,^15,16,17,18^ suggested by the decline in low-income, rural, and less educated patients during Phase 1 – highlighting the importance of concerted efforts to combat worsening health inequities during the most resource-constrained times. The phases extending past vaccine availability also reveal a surprisingly persistent lag in return to pre-pandemic volumes, suggesting that the pandemic has had lasting impacts on patient healthcare seeking practices.

Interestingly, analysis of all COVID phases combined showed no significant difference in likelihood of receiving a procedure after NSTEMI diagnosis, suggesting that procedural care declined proportionally to the decreased volume of NSTEMI presentations overall. However, when individual phases were analyzed, they revealed an isolated finding of a lower likelihood of procedural treatment during the acute Phase 1 and a higher likelihood during the recovery Phase 6. While this did not appear to translate to an increase in mortality risk during Phase 1, a mortality benefit was observed in Phase 6 compared to pre-COVID. The improved outcomes in Phase 6 hopefully herald a return to normalcy post-pandemic, though further research is required to determine the degree to which this is due to the rise procedure use versus advancements in cardiovascular care over the four year period studied. On the other hand, increased procedure use in Phase 6 may also be the earliest sign of appropriate treatment for a resurgence of untreated disease that was neglected during the pandemic years.

Examination of Kaplan-Meier survival curves reveals that mortality after NSTEMI was worse across Phases 2 (community spread), 3 (first peak), and 5 (second peak) compared to pre-pandemic. Adjusted regression analyses of 30-day mortality show that patients in Phase 2 and 3 experienced worse outcomes, but these were no longer significant by Phase 5 – almost 2 years into the pandemic – suggesting that cardiovascular systems adapted over time to the new paradigm of pandemic care, despite the much higher burden of COVID-19 infections during this time.

Closer examination of the circumstances of Phases 2-3 reveal the unique vulnerabilities during this time period. As rapid early growth in COVID infections climbed, this was a time of great uncertainty, during which strict national mandates limiting elective care started to be repealed and providers had little prior data to guide decisions on how to resume care.^19,20^ Analysis of the mortality outcomes after adjustment for receipt of PCI in these phases showed negligible effect, suggesting appropriate triage of procedural care. This points to the need to examine other potential contributing factors, such as inpatient care practices, outpatient follow-up, transition to telehealth, and medication compliance.

Our findings that Veterans increased their use of non-VA care during the pandemic likely stem from multiple etiologies. Firstly, progressive implementation of the Mission Act over the course of the pandemic may have impacted Fee-Basis usage.^21^ However, recovery of VA prescription volumes in early Phase 2 of the pandemic argues against an overall decline in VA utilization as the sole cause. Contagion concern may have led patients to seek the closest care possible, or limited patient capacity to travel if the VA was not the nearest hospital. Severe VA bed shortages and strict VA rules limiting non-essential procedures may have also driven care to non-VA hospitals.^22^ Pre-pandemic studies have suggested improved outcomes for MI care within the VA,^22,23^ and we similarly found lower unadjusted risk of mortality among patients treated at VA facilities compared to non-VA facilities across all pre- and post-pandemic phases. After adjustment however, the differences became no longer significant in the pre-pandemic period, as well as Phases 1, 2, and 5, leaving only Phases 3, 4, and 6 with significantly higher mortality risk at non-VA facilities. Interestingly, the higher non-VA facility mortality found in Phase 3 overlaps with the higher mortality observed within the VA alone in Phase 3 compared to pre-pandemic – suggesting possible healthcare-system wide vulnerabilities during this phase.

This study has limitations. While ICD coding for STEMI and NSTEMI has been previously validated,^24^ non-COVID-19 coding reliability during the pandemic has not been specifically addressed. One of the advantages of VA data is that it avoids bias from voluntary reporting or insurance status,^25^ however, it is a predominantly male population. We adjusted our analyses for multiple patient characteristics, though additional confounders may exist. This data does not capture patients who did not present to a healthcare facility.

In conclusion, we found that the VA healthcare system responded to the COVID-19 pandemic with reductions in procedural treatment for NSTEMI that were proportional to the lower numbers of patients presenting with NSTEMI, yet the community spread and first peak of the pandemic was still impacted with higher mortality. These resolved by the second, higher pandemic peak, but reveal the vulnerabilities that occurred during earlier abrupt changes in operational capacity and serve as a guide to direct preparations for future threats.

## Data Availability

Data in the manuscript is available upon reasonable request to the corresponding author.

## Acknowledgments

n/a

